# Improved Complexity Stratification in Congenital Heart Disease; the Impact of Including Procedural Data on Accuracy and Reliability

**DOI:** 10.1101/2024.01.31.24302110

**Authors:** Jason Chami, Calum Nicholson, David Baker, Rachael Cordina, Geoff Strange, David S. Celermajer

## Abstract

**Introduction:** In order to manage a class of diseases as broad as congenital heart disease (CHD), multiple “manually generated” classification systems defining CHDs as mild, moderate and severe have been developed and used to good effect. As databases have grown, however, such “manual” complexity scoring has become infeasible. Though past attempts have been made to determine CHD complexity algorithmically using a list of diagnoses alone, missing data and lack of procedural information have been significant limitations.

**Methods:** We built an algorithm that can stratify the complexity of patients with CHD by integrating their diagnoses with a list of their previous procedures. Specific procedures which address a missing diagnosis or imply a certain operative status were used to supplement the diagnosis list. To verify this algorithm, CHD specialists manually checked the classification of 100 children and 100 adults across four hospitals in Australia.

**Results:** Our algorithm was 99.5% accurate in the manually checked cohort (100% in children and 99% in adults), and was able to automatically classify more than 90% of a cohort of over 24,000 CHD patients, including 92.5% of children (vs 84.4% without procedures, p < 0.0001) and 91.1% of adults (vs 70.4% without procedures; p < 0.0001).

**Conclusions:** CHD complexity scoring is significantly improved by access to procedural history and can be automatically calculated with high accuracy.

## 1 Introduction

Congenital heart diseases (CHDs) are many and varied, existing on a vast spectrum from mild nuisance to severe life-altering illness. To aid in the management of such a broad class of diseases, multiple classification systems separating CHDs into mild, moderate and severe disease have been developed, originally based on local empirical data and expert consensus. These classification systems, requiring manual coding, soon developed and formalized,^1,2^ coming into their own as invaluable research tools that allow researchers to group diseases that are individually rare but collectively common in order to boost statistical power—an absolutely essential task, even in very large CHD databases. While these guidelines were initially intended to be applied on a patient-by-patient basis by experts familiar with the patient’s entire clinical and surgical history, this is prohibitively time-consuming for very large contemporary databases, especially as guidelines are regularly adjusted in response to clinical research and new surgical techniques.

To address this issue in our own Congenital Heart Alliance of Australia and New Zealand (CHAANZ) Registry, a large database which aims to collect information from all CHD patients in Australia and New Zealand, our group developed and verified an algorithm that could assign European Society of Cardiology (ESC) complexity scores^3^ to most patients based solely on a list of diagnoses.^4^

While this algorithm has been very useful in our database for the generation of summary statistics and the selection of stratified samples for further research, it has two fundamental limitations. The first limitation is missing data. Many patients known personally to specialists in the research team are noted to lack complete diagnostic codes—especially pediatric patients whose data is mainly stored in surgical, rather than clinical databases. The second limitation is subtler; the ESC classifies many diagnoses differently based on whether they have been repaired/palliated or not, so without this information a large number of patients cannot be confidently classified, especially those with transposition of the great arteries (TGA), Tetralogy of Fallot (TOF), atrial septal defect (ASD) and ventricular septal defect (VSD).

Our review of the world’s largest CHD databases^5^ showed that most, including the Belgian Congenital Heart Disease Database^6^ and the Dutch CONCOR Database,^7^ already contain procedural information. The CHAANZ Registry is no exception, with surgical and procedural data recently added from two children’s hospitals and two expert adult CHD (ACHD) centers. In this context, it was time to address the limitations of a diagnosis-only classification algorithm by upgrading ours to automatically integrate clinical and procedural history to generate an accurate patient complexity classification. Our aim is to present this novel algorithm, and in doing so to demonstrate the power of algorithmically combining diagnostic, clinical and procedural information, often individually sequestered, to gain significantly enhanced insights, at negligible computational or administrative cost.

## 2 Methods

### 2.1 Ethics and Consent

We performed this study to facilitate improvements to the CHAANZ CHD Registry and its ability to classify CHD complexity, extending previous work from our research group which presented a classification algorithm using diagnoses alone.^4^ Ethics approval for the CHAANZ CHD Registry has been granted by the Human Research Ethics Committee at Royal Prince Alfred Hospital (2019/ETH07472). A waiver of consent is in place for the analysis of retrospective data.

### 2.2 Building the Algorithm

#### 2.2.1 Principles

We recently published a classification algorithm based on diagnoses alone;^4^ our new algorithm is built on this base. In the previous algorithm, each diagnosis begins by being assigned a provisional complexity. These provisional complexities are then updated based on patient comorbidities. Finally, the patient is assigned the maximum of their diagnosis codes.

Procedures inform the complexity scores in two new ways: firstly, we use the fact that some procedures imply particular diagnoses; for example, a Fontan procedure implies that the patient has a functionally univentricular heart. We can use this to fill in gaps in a sparse diagnosis list, thus gaining a more complete picture of the patient. Where procedures imply multiple comorbid diagnoses, they can all be added to the diagnosis list. Where procedures do not have a clear implied diagnosis, however, this strategy cannot be used. For patients who have undergone certain advanced surgeries that are used for multiple severe CHDs, our algorithm adds an implied diagnosis of “Complex CHD” for that patient.

The second way that procedures enrich the classification process is by indicating when certain lesions have been repaired or not; for example, TGA treated with an arterial switch procedure has a moderate complexity, while unrepaired TGA is severe. Operative status is a major contributor to disease severity as per the ESC guidelines, especially for common lesions including ASD, VSD and PDA, and this was a major limitation of our previously published “diagnosis-only” algorithm.

Using these two strategies allows us to fill in missing diagnoses and clarify existing ones, adding robustness by merging separately gathered diagnostic and procedural data.

#### 2.2.2 “Unknowns”

If the algorithm is not able to classify a certain patient’s complexity with the information provided, it will classify that patient as “unknown”. While this outcome is not ideal, it is important that the algorithm not make classifications where insufficient information exists to do so, for example where the size or operative status of a particular lesion is unknown. The ideal algorithm should agree with an expert clinician that particular patients are of unknown complexity, and flag such patients for more detailed manual review.

#### 2.2.3 Complications and Operative Status

The classification of ASD, VSD and PDA depends in part on the type of defect, severity of the lesion, complications (such as the presence of pulmonary hypertension and/or valve regurgitations) and pre-versus post-operative status. We also searched for codes specifically designed for procedural sequelae, but these were rarely used. Though the ESC mentions chamber enlargement, ventricular dysfunction and pulmonary hypertension as potential complications, the lack of EPCC codes for these conditions means our algorithm can only take the latter into account.

#### 2.2.4 The New Algorithm: In Summary

For a graphical representation of the new algorithm in action, see Figure 1. The first algorithm input is a list of European Pediatric Cardiac Code^8^ (EPCC) diagnostic codes, each linked to a patient ID. Next, a list of procedures in the same format is joined to a procedure dictionary, which matches applicable procedure codes to implied diagnoses, including “repaired” diagnoses modified from the standard EPCC by the addition of an “r” (e.g. 09.21.21r: Repaired PDA). The compiled list of diagnoses (listed, implied and repaired) is compared to a complexity table, which lists possible complexity scores for each diagnosis. From these, using rules adapted from the ESC guidelines, the final patient complexity classification can be determined. The algorithm in its entirety is included in the Supplement.

**Figure 1.**
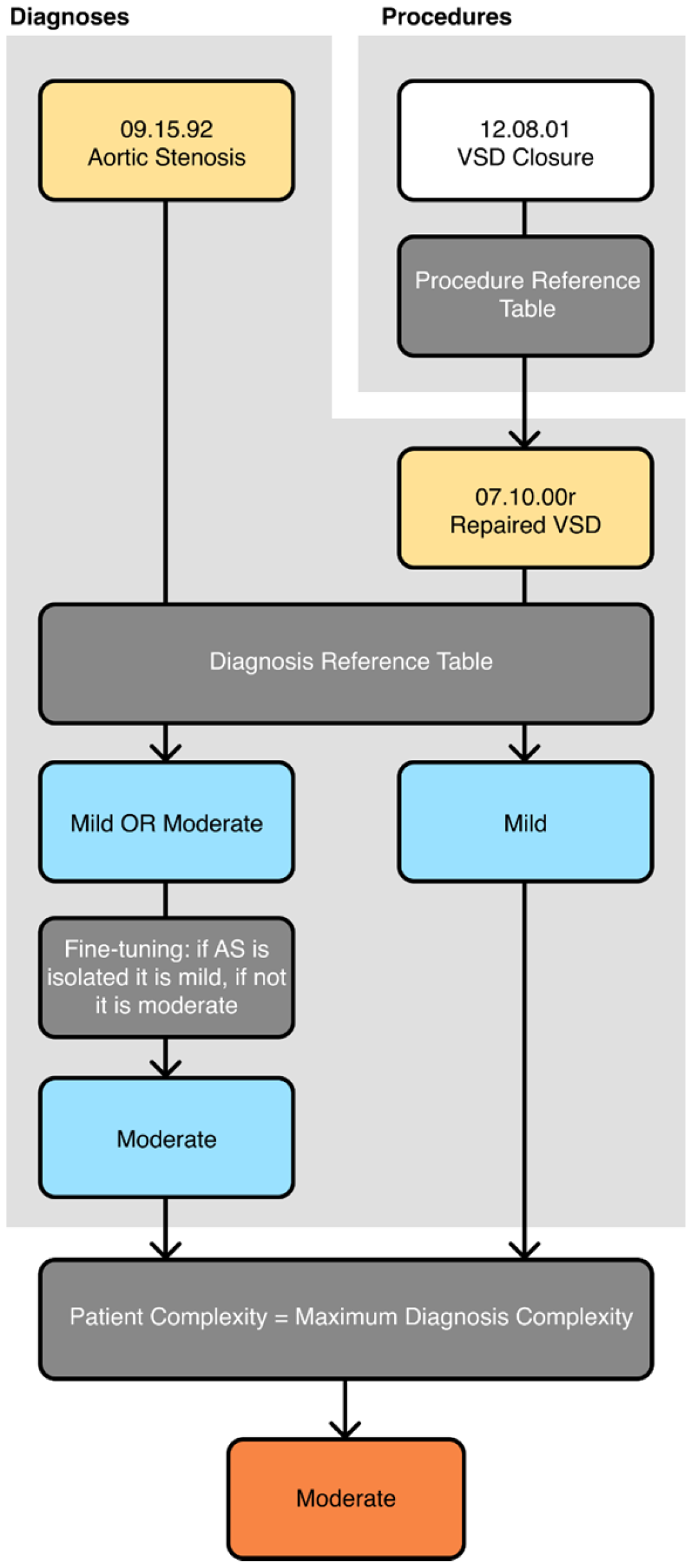
Flowchart showing the action of the algorithm on an example patient with one listed diagnosis, aortic stenosis (AS), and one listed procedure, ventricular septal defect (VSD) closure. The VSD closure procedure implies that the patient has a repaired VSD that was missing from the diagnosis list. The patient’s AS is not, therefore, isolated. Thus, the patient receives a “moderate” classification, with the new algorithm.

### 2.3 Evaluating the Algorithm

#### 2.3.1 Participating Sites

We used data from four large quaternary CHD treatment centers. The two pediatric sites used were the Children’s Hospital at Westmead, Sydney (CHW, n = 8745) and the Women’s and Children’s Hospital in Adelaide (WCH; n = 8951). The two adult CHD sites used were the Royal Melbourne Hospital (RMH; n = 2892) and the Prince Charles Hospital, Brisbane (TPCH; n = 4070). In total, 24658 patients were under analysis. We ran the algorithm on all patients in order to determine the coverage, i.e. the percentage of patients who can be confidently given a classification (not including “unknown”).

#### 2.3.2 Validation

Using the CHAANZ CHD Registry we randomly selected 100 children from CHW and WCH, and 100 adults from RMH and TPCH to undergo manual complexity classification by a CHD specialist using ESC guidelines. This “gold-standard” classification was compared to the algorithmic classification. The difference in classification rate was tested for statistical significance using χ^2^-tests on the pediatric, adult, and overall cohorts.

## 3 Results

### 3.1 Children

Out of 17696 pediatric patients across CHW (n = 8745) and WCH (n = 8951), our algorithm was able to confidently classify 16370 patients (92.5%), and was uncertain for 1326 patients (7.5%). Without procedural data, 14292 patients could be confidently classified (84.4%), and 2767 were unknown (15.6%). Therefore, our algorithm was able to classify an additional 1441 pediatric patients (8.1%) by utilizing procedural data, taking the total coverage from 84.4% to 92.5% (p < 0.0001). Put another way, out of the unknowns from the previous algorithm, an extra 1441 of 2767 could now be classified, for an “improvement” rate of over 50% of previously unclassifiable patients.

One hundred patients were randomly selected from CHW (n = 79) and WCH (n = 21) for manual classification by an expert CHD clinician. The algorithm agreed with the expert clinician assessment in 100% of the selected cases (Figure 2A). There were 15 mild cases, 53 moderate, 23 severe and 9 unknown. The most common diagnoses in this cohort were patent foramen ovale (PFO; n = 34), ASD (n = 31), mitral regurgitation (n = 31), PDA (n = 26) and tricuspid regurgitation (n = 26; note that patients often had multiple diagnoses).

**Figure 2.**
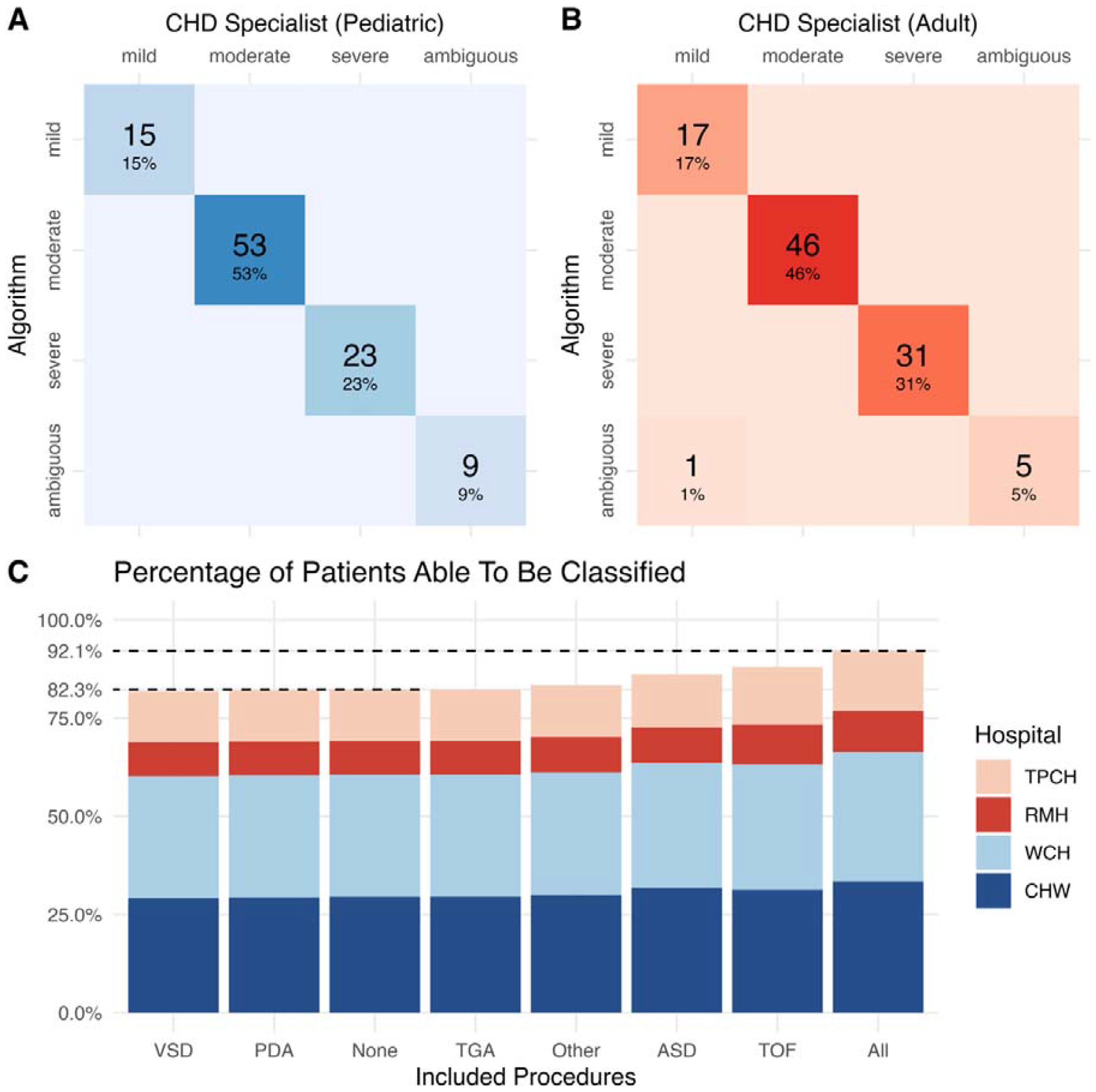
**A)** Confusion matrix showing 100% agreement between the algorithm and the CHD specialist on a subset of 100 pediatric patients. **B)** Confusion matrix showing 99% agreement between the algorithm and the CHD specialist on a subset of 100 adult patients. **C)** Graph showing the percentage of all 24658 patients able to be classified by the algorithm, when it is modified to also include procedures related to certain diagnoses. Patients come from four Australian hospitals: two children’s hospitals in blues, and two adult CHD centers in reds. **TPCH** The Prince Charles Hospital; **RMH** The Royal Melbourne Hospital; **WCH** The Women’s and Children’s Hospital; **CHW** The Children’s Hospital at Westmead.

### 3.2 Adults

Out of 6962 adult patients across RMH (n = 2892) and TPCH (n = 4070), our algorithm was able to confidently classify 6339 patients (91.1%) and was uncertain for 623 patients (8.9%). Without procedural data, 4898 patients could be confidently classified (70.4%) and 2064 were unknown (29.6%). Therefore, our algorithm was able to classify an additional 1441 adult patients (20.7%) by utilizing procedural data, taking the total coverage from 70.4% to 91.1% (p < 0.0001). Therefore, an extra 1441 out of the 2064 patients marked as unknown by the previous algorithm could now be classified by incorporating procedural data, for an effective improvement rate of 70% of previously unclassifiable patients.

One hundred patients were randomly selected from RMH (n = 32) and TPCH (n = 68) for manual classification by an expert CHD clinician. The algorithm agreed with expert clinician assessment in 99% of the selected cases (Figure 2B). In one case, a patient had mistakenly received a diagnosis code for both an ASD (EPCC 05.04.02) and a PFO (EPCC 05.03.01). Since the ASD was neither repaired nor isolated, the algorithm classified it as “unknown” complexity. In reality, this was an input error; the patient should have only had a diagnosis of PFO, and should have been classified as mild. There were 18 mild cases, 46 moderate, 31 severe and 5 unknown, but the algorithm misclassified one mild case as unknown.

### 3.3 Overall Results

Overall, 92.1% of the cohort was able to be classified using our newer algorithm, compared to 82.3% without using procedures (p < 0.0001; Figure 1C). By adding groups of procedures into the algorithm one at a time we were able to determine which procedures had the greatest effect in increasing the classifiable proportion of patients. Overall, the inclusion of TOF procedures led to the greatest increase in the number of classifiable patients, followed by ASD procedures, other procedures (excluding ASD, VSD, PDA, TGA and TOF) and TGA procedures (Figure 1C).

## 4 Discussion

Our novel and improved automated CHD classification algorithm, which now uses both diagnoses and procedural data, was able to confidently classify 92.5% of children and 91.1% of adults with CHD according to ESC guidelines, representing a substantial and significant improvement on our previously published algorithm, which used diagnoses only. The algorithm produced the same classification as an expert CHD clinician in 199 out of 200 manually classified patients, in a fraction of the time. This shows that diagnostic and procedural information can be algorithmically combined to generate clinically relevant insights with unprecedented accuracy and coverage.

This algorithm could therefore be used to instantly calculate ESC complexity for patients in the world’s largest CHD databases, most of which already include procedural information.^5–7^ This could be used in research, for example, where having comprehensive complexity scoring allows for stratified sampling (as is currently being performed in the CHAANZ database), and can be used to group diagnoses together in order to generate sufficient statistical power—a vital function given the rarity and heterogeneity of CHDs. In large databases, manual classification is infeasible; this algorithm can be used instead, and easily modified as guidelines are updated.

Despite significant improvements in coverage and accuracy, just under 10% of patients remained unclassifiable by this novel algorithm. Much of this, however, reflects the inherent impossibility of classification when the ESC guidelines require an understanding of the size of a particular lesion,^2^ and also the generally suboptimal state of CHD data quality in hospital records and on death certificates.^9,10^ It also reflects a limitation in the EPCC, which lacks different codes for “small” and “large” lesions, making it impossible to algorithmically classify most unoperated ASDs, VSDs and PDAs.^8^ Despite these limitations, the ability of the algorithm to accurately classify over 90% of patients means that less than 10% would require manual coding for the dataset to be complete, potentially saving a large amount of time. Although data quality and comprehensiveness remain the limiting factors in improving accuracy and coverage, our use of procedures to “imply” diagnoses both improves coverage and adds a layer of robustness, as procedural data is usually entered and verified separately to diagnostic data.

Future improvement in the algorithmic complexity classification of CHD patients is now largely dependent on improvements in the classification and coding systems themselves, and in the quality of clinical, diagnostic and procedural data collection and recording. As clinical databases like CHAANZ continue to grow, there may be scope in the future to expand complexity scoring algorithms to include functional capacity and other physiologic markers, to gain a fuller picture of a patient’s risk profile.^11^ Despite current limitations, however, this algorithm is now able to confidently classify more than 90% of a very large sample of children and adults with CHD across Australia, with near-perfect accuracy when compared to manual classification by a CHD specialist. It is our hope that this algorithm will allow researchers working with CHD databases of all sizes to make use of valuable complexity scoring, at a fraction of the effort and cost of manual classification.

## Data Availability

This submission contains our complexity coding algorithm in the R programming language which can be tested and verified at https://github.com/JasonChami/CHD-Procedure-Complexity

## 5 Acknowledgements

None.

## References

1. Warnes, C. A. et al. Task Force 1: the changing profile of congenital heart disease in adult life. J. Am. Coll. Cardiol. 37, 1170–1175 (2001).

2. Baumgartner, H. et al. 2020 ESC Guidelines for the management of adult congenital heart disease: The Task Force for the management of adult congenital heart disease of the European Society of Cardiology (ESC). Endorsed by: Association for European Paediatric and Congenital Cardiology (AEPC), International Society for Adult Congenital Heart Disease (ISACHD). Eur. Heart J. 42, 563–645 (2021).

3. Baumgartner, H. et al. 2020 ESC Guidelines for the management of adult congenital heart disease. Eur. Heart J. 42, 563–645 (2021).

4. Chami, J. et al. Algorithmic complexity stratification for congenital heart disease patients. Int. J. Cardiol. Congenit. Heart Dis. 11, 100430 (2023).

5. Chami, J., Nicholson, C., Strange, G., Cordina, R. & Celermajer, D. S. National and regional registries for congenital heart diseases: Strengths, weaknesses and opportunities. Int. J. Cardiol. 338, 89–94 (2021).

6. Ombelet, F. et al. Creating the BELgian COngenital heart disease database combining administrative and clinical data (BELCODAC): Rationale, design and methodology. Int. J. Cardiol. 316, 72–78 (2020).

7. Vander Velde, E. T. et al. CONCOR, an initiative towards a national registry and DNA-bank of patients with congenital heart disease in the Netherlands: Rationale, design, and first results. Eur. J. Epidemiol. 20, 549–557 (2005).

8. Franklin, R. The European Paediatric Cardiac Code Long List: Structure and function — the first revision. Cardiol. Young - CARDIOL YOUNG 12, (2002).

9. Chami, J. et al. High Error Rates in Coding Causes of Death in Adults With Congenital Heart Disease. JACC Adv. 1, 1–2 (2022).

10. Chami, J. et al. Hospital discharge codes and substantial underreporting of congenital heart disease. Int. J. Cardiol. Congenit. Heart Dis. 7, 100320 (2022).

11. Ombelet, F., Goossens, E., Van De Bruaene, A., Budts, W. & Moons, P. Newly Developed Adult Congenital Heart Disease Anatomic and Physiological Classification: First Predictive Validity Evaluation. J. Am. Heart Assoc. 9, (2020).

